# IgG antibody responses are preferential compared to IgM for use as serological markers for detecting recent exposure to *Plasmodium vivax* infection

**DOI:** 10.1101/2020.08.07.20169862

**Authors:** Rhea J Longley, Michael T White, Jessica Brewster, Zoe SJ Liu, Caitlin Bourke, Eizo Takashima, Matthias Harbers, Wai-Hong Tham, Julie Healer, Chetan E Chitnis, Wuelton Monteiro, Marcus Lacerda, Jetsumon Sattabongkot, Takafumi Tsuboi, Ivo Mueller

## Abstract

To achieve malaria elimination, new tools are required to explicitly target *Plasmodium vivax*. Recently, a novel panel of *P. vivax* proteins were identified and validated as serological markers for detecting recent exposure to *P. vivax* within the last 9 months. In order to improve the sensitivity and specificity of these markers, IgM in addition to IgG antibody responses were assessed to a down-selected panel of 20 *P. vivax* proteins. IgM was tested using archival plasma samples from observational cohort studies conducted in malaria-endemic regions of Thailand and Brazil. IgM responses to these proteins generally had poorer classification performance than IgG.

## Background

Infections due to *Plasmodium vivax* are a major challenge for malaria elimination. This is due to unique biological features of *P. vivax* parasites, including an arrested stage in the liver (hypnozoites) that can reactivate weeks to months later causing relapses of clinical disease. Individuals with hypnozoites are major reservoirs of transmission, and are responsible for over 80% of blood-stage *P. vivax* infections [1]. Another challenge is the high proportion of low-density, asymptomatic blood-stage infections due to *P. vivax* [2], particularly in low-transmission regions. These factors make it difficult to identify not only infected individuals but also delineate pockets of ongoing, local transmission. It is therefore critical that novel tools are developed that enable efficient identification of at-risk individuals that should be targeted for malaria interventions.

We have recently identified and validated a novel panel of *P. vivax* proteins that induce IgG antibody responses reflective of recent exposure to *P. vivax* blood-stage infections [3]. Combinations of IgG antibody responses to 5-8 *P. vivax* proteins can accurately classify (with 80% sensitivity and specificity) whether an individual has had a *P. vivax* infection within the last 9 months. We chose this 9-month time frame as individuals who have had a detectable blood-stage infection in this period, and have not been treated with anti-liver-stage drugs, are likely to be harbouring hypnozoites in their livers. Our novel serological exposure markers therefore represent the first test that can, indirectly, identify hypnozoite carriers. This tool could play an important role in malaria elimination by offering an alternative to mass drug administration strategies (MDA, where everyone is treated), or mass screening and treatment (MSAT) strategies performed using currently available diagnostics for blood-stage parasites. Whilst effective, MDA results in a high level of overtreatment. Conversely, MSAT is ineffective using currently available technologies [4]. We have proposed an alternative strategy termed “serological testing and treatment”, whereby individuals are tested using our serological exposure markers and treatment given to those exposed during the past 9 months.

Here, we investigate the potential utility of alternative biomarkers to IgG antibody responses as serological exposure markers. We previously observed that IgG responses to different *P. vivax* proteins were highly correlated [3], and this is likely why we were unable to improve the classification accuracy by simply incorporating responses to more antigens into the algorithm. IgM antibody responses to the same *P. vivax* protein are only weakly correlated to IgG [5] and are generally thought to have a different response kinetic (as shown against *P. falciparum* malaria [6] and other infectious diseases such as West Nile virus [7]). We thus hypothesized that IgM antibody responses to our panel of *P. vivax* proteins could be used to improve the classification accuracy by providing additional information into the algorithm.

## Methods

We tested our hypothesis using samples from two observational cohort studies conducted over 2013-2014: one in the Kanchanaburi and Ratchaburi provinces of western Thailand [8] and one in Manaus in the Brazilian Amazon [3]. We utilized plasma samples available from the last visits of these cohorts (n = 829 Thailand, n = 925 Brazil), as previously described [3]. After enrolment individuals were sampled every month over the yearlong cohort, with 13-14 active case detection visits performed (with PCR-based detection of malaria infections). This enabled us to relate IgM (or IgG) antibody levels measured at the last visit with time since previous detected *P. vivax* infection. All individuals provided informed consent or assent, and the studies were approved locally by the Ethics Committee of the Faculty of Tropical Medicine, Mahidol University, Thailand (MUTM 2013-027-01), and the Brazilian National Committee of Ethics (CONEP) (349.211/2013). We also utilized three panels of malaria-naïve control plasma samples as previously described [9]: 102 samples from the Volunteer Blood Donor Registry (VBDR), Melbourne, Australia, 100 samples from the Australian Red Cross (ARC), Melbourne, Australia and 72 samples from the Thai Red Cross (TRC), Bangkok, Thailand. The Human Research Ethics Committee at WEHI approved usage of all samples at WEHI, and collection of the malaria-naïve control samples (#14/02).

IgM antibody responses were measured against a panel of 18 or 20 *P. vivax* proteins in samples from the Thai or Brazilian cohorts, respectively (see Table S1 for full list of proteins, expression and purification method, sequence region). These proteins were selected as they were the best performing when using IgG responses in the first iteration of our algorithm [9]. The *P. vivax* proteins were coupled to non-magnetic COOH microspheres as previously described [10], and IgM levels measured using a modified multiplexed Luminex® assay [9]. Modifications were plasma samples diluted to 1/200 (instead of 1/100 for IgG), and use of the secondary donkey F(ab’)2 anti-human IgM Fc_5_, Jackson ImmunoResearch Laboratories, Inc., at 1/400 dilution. Median fluorescent intensity (MFI) values from the Luminex®-200 were converted to relative antibody units based on a standard curve run on each plate generated from a positive control plasma pool consisting of highly immune adults from Papua New Guinea [10]. Data for KMZ83376.1 and PVX_095055 were not tested/analysed for the Thai cohort due to inconsistent standard curves. IgG antibody responses against the same *P. vivax* proteins had previously been measured in all samples as described [3].

Individuals from the malaria-endemic cohorts were defined as either i) infected with *P. vivax* within the last 9 months prior to antibody measurements or ii) not infected with *P. vivax* within the last 9 months. Individuals from the malaria-naïve control panels were defined in the latter group. Single antigen and two-antigen linear discriminant analysis (LDA) classifications were performed in R studio using R version 3.5.3 [11] and the packages MASS [12] and ROCR [13].

## Results

We first determined the accuracy for classifying individuals in the Thai and Brazilian cohorts as recently infected within the last 9 months using IgM antibody responses against 18 or 20 *P. vivax* proteins (Figure 1), respectively. Overall, we observed lower levels of classification accuracy with IgM to these proteins as compared to using IgG [3], as shown in Figure 1 by reference to the top performing serological exposure marker for IgG (RBP2b). The area under the curve (AUC) values for IgM ranged from 0.55 - 0.77 for the Thai cohort and 0.50 - 0.72 for the Brazilian cohort (Table 1). In comparison, the AUC values for IgG for the same proteins ranged from 0.70 - 0.85 for the Thai cohort and 0.65 - 0.82 for the Brazilian cohort (Table S2) (note these AUC values are different to that in our prior publication [3] as the current analysis was performed with negative controls from Melbourne and Bangkok only, not including newer samples from Rio de Janeiro). For IgM, the top performing *P. vivax* protein in both cohorts was PVX_087885B, annotated as the rhoptry-associated membrane antigen (RAMA, putative). One other protein PVX_082735 (thrombospondin-related anonymous protein, TRAP) performed well with IgM in both the Thai and Brazilian cohorts with AUC values of 0.74 and 0.71, respectively. In the Thai cohort, the protein PVX_082670 (merozoite surface protein 7 putative, MSP7) also performed reasonably well for IgM (AUC 0.75). The AUC for IgM responses against RBP2b (top performing marker for IgG) were much lower at 0.63 and 0.56. IgM antibody responses, stratified by time since previous detected *P. vivax* infection by PCR, are shown in Figures S1 and S2.

**Figure 1:**
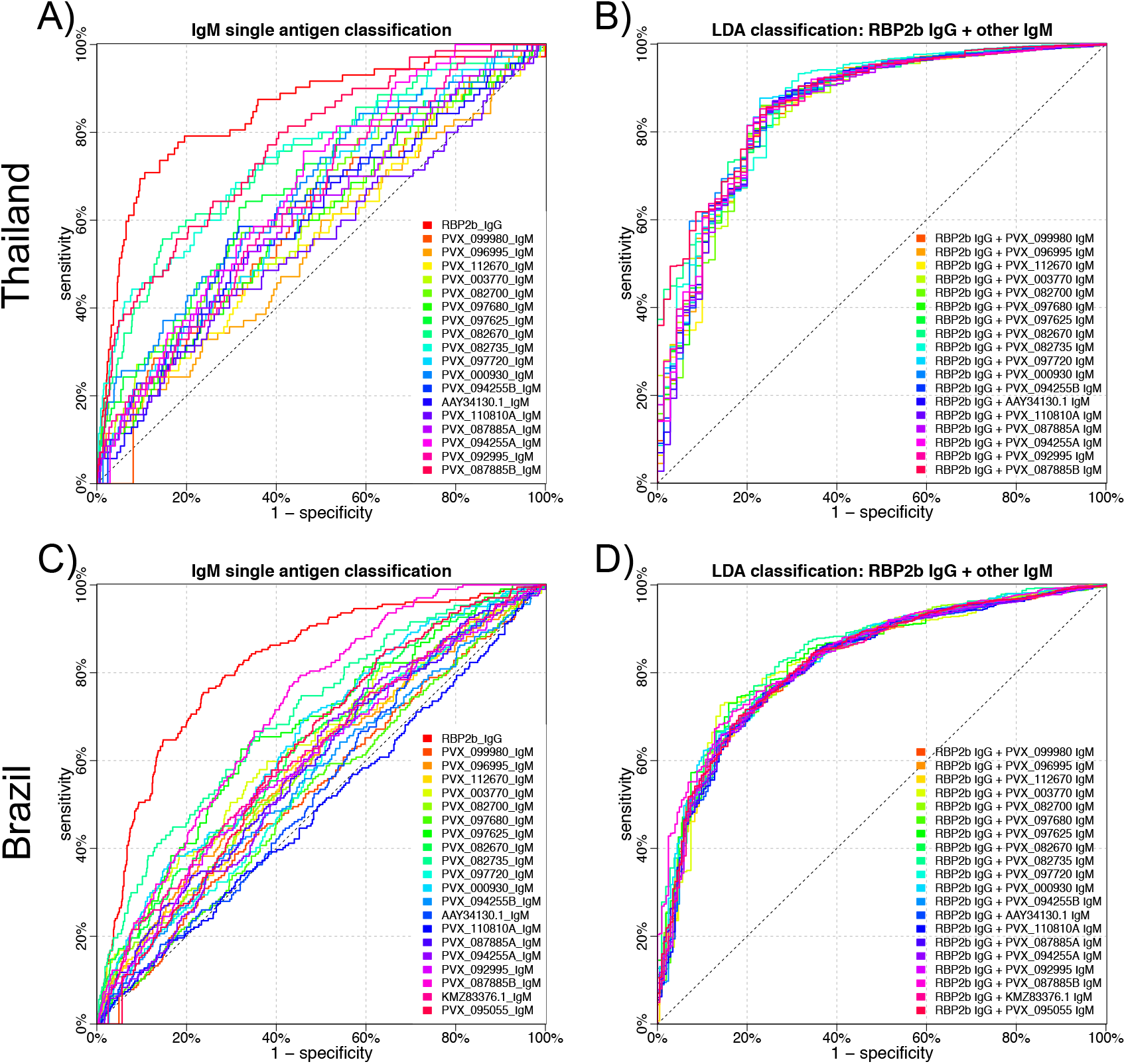
Performance of IgM antibodies against 18-20 *P. vivax* proteins individually (A and C) and in combination with the top performing protein using IgG (B and D), for classification of *P. vivax* infections within the prior 9 months. In the Thai cohort, IgM antibody responses were measured against 18 *P. vivax* proteins. Panel A depicts the classification accuracy of these 18 proteins individually, and includes the top performing protein for IgG (RBP2b) as reference in red. Panel B depicts the results from a LDA combining RBP2b IgG with each of the IgM responses to the 18 proteins. In the Brazilian cohort, IgM antibody responses were measured against 20 *P. vivax* proteins. Panel A depicts the classification accuracy of these 20 proteins individually, and includes the top performing protein for IgG (RBP2b) as reference in red. Panel B depicts the results from a LDA combining RBP2b IgG with each of the IgM responses to the 20 proteins. AUC values are shown in Table 1.

**Table 1:** AUC values for classifying individuals as recently infected with *P. vivax*. LDA (2 antigen combination) was performed using the 9 month classification period with RBP2b IgG plus IgM to one of the listed antigens. NA = not applicable. NT = not tested.

|  | Single antigen classification |  |  |  |  |  |  |  | LDA |  |
| --- | --- | --- | --- | --- | --- | --- | --- | --- | --- | --- |
|  | 1 month |  | 3 months |  | 6 months |  | 9 months |  | 9 months |  |
| Protein | *Thai* | *Brazil* | *Thai* | *Brazil* | *Thai* | *Brazil* | *Thai* | *Brazil* | *Thai* | *Brazil* |
| RBP2b (IgG) | 0.825 | 0.880 | 0.856 | 0.823 | 0.857 | 0.823 | 0.849 | 0.818 | NA | NA |
| PVX_099980 | 0.621 | 0.516 | 0.601 | 0.517 | 0.606 | 0.529 | 0.607 | 0.537 | 0.855 | 0.822 |
| PVX_096995 | 0.551 | 0.625 | 0.537 | 0.591 | 0.548 | 0.597 | 0.549 | 0.593 | 0.853 | 0.822 |
| PVX_112670 | 0.595 | 0.590 | 0.579 | 0.600 | 0.585 | 0.594 | 0.567 | 0.589 | 0.848 | 0.823 |
| PVX_003770 | 0.613 | 0.734 | 0.573 | 0.660 | 0.587 | 0.644 | 0.602 | 0.633 | 0.852 | 0.827 |
| PVX_082700 | 0.665 | 0.570 | 0.653 | 0.613 | 0.662 | 0.602 | 0.641 | 0.596 | 0.849 | 0.828 |
| PVX_097680 | 0.590 | 0.551 | 0.577 | 0.544 | 0.593 | 0.535 | 0.593 | 0.529 | 0.851 | 0.819 |
| PVX_097625 | 0.664 | 0.680 | 0.647 | 0.663 | 0.655 | 0.655 | 0.671 | 0.665 | 0.856 | 0.835 |
| PVX_082670 | 0.786 | 0.617 | 0.745 | 0.650 | 0.742 | 0.639 | 0.747 | 0.635 | 0.871 | 0.827 |
| PVX_082735 | 0.786 | 0.774 | 0.766 | 0.730 | 0.758 | 0.716 | 0.743 | 0.705 | 0.863 | 0.845 |
| PVX_097720 | 0.655 | 0.586 | 0.629 | 0.572 | 0.632 | 0.568 | 0.628 | 0.573 | 0.858 | 0.821 |
| PVX_000930 | 0.709 | 0.614 | 0.686 | 0.634 | 0.688 | 0.649 | 0.673 | 0.654 | 0.863 | 0.835 |
| PVX_094255B | 0.628 | 0.577 | 0.617 | 0.545 | 0.626 | 0.562 | 0.631 | 0.560 | 0.851 | 0.819 |
| AAY34130.1 | 0.622 | 0.518 | 0.618 | 0.519 | 0.617 | 0.541 | 0.597 | 0.540 | 0.851 | 0.819 |
| PVX_110810A | 0.580 | 0.505 | 0.581 | 0.475 | 0.585 | 0.493 | 0.554 | 0.496 | 0.847 | 0.818 |
| PVX_087885A | 0.628 | 0.636 | 0.611 | 0.602 | 0.625 | 0.591 | 0.615 | 0.591 | 0.851 | 0.823 |
| PVX_094255A | 0.673 | 0.534 | 0.669 | 0.555 | 0.676 | 0.570 | 0.675 | 0.581 | 0.856 | 0.821 |
| PVX_092995 | 0.639 | 0.584 | 0.629 | 0.601 | 0.644 | 0.595 | 0.640 | 0.599 | 0.858 | 0.823 |
| PVX_087885B | 0.799 | 0.733 | 0.760 | 0.725 | 0.771 | 0.717 | 0.771 | 0.715 | 0.876 | 0.843 |
| KMZ83376.1 | NT | 0.623 | NT | 0.594 | NT | 0.612 | NT | 0.617 | NT | 0.822 |
| PVX_095055 | NT | 0.616 | NT | 0.607 | NT | 0.614 | NT | 0.624 | NT | 0.827 |

As IgM responses are expected to decay more quickly than IgG (due to a shorter serum half-life and their characterization as an early response to infection), we hypothesized that IgM responses may be better markers of very recent exposure. We therefore tested the ability of IgM antibody responses to our 18-20 *P. vivax* proteins to classify individuals as infected with *P. vivax* within the prior 6, 3 or 1-month period. As shown in Table 1, for both cohorts, the AUC values did improve when using a shorter time frame for classification. However, the same proteins consistently performed well (PVX_087885, PVX_082735 and PVX_082670) with all classification time frames tested, and the overall improvements were marginal. Furthermore, classification performance with IgM antibodies was still inferior to classification with RBP2b IgG.

Our original results had shown that IgG antibody responses to combinations of proteins were better at classifying individuals as infected within the last 9 months compared to individual proteins alone. We therefore tested the classification ability, using the 9 month time frame, of each IgM antibody response combined with IgG responses against the top protein RBP2b. Each of the two-antigen combinations (1 IgM with RBP2b IgG) had better classification accuracy than the IgM response alone, as determined by AUC values (Table 1). In the Thai cohort, all IgM + RBP2b IgG performed (slightly) better than RBP2b IgG alone, with the exception of two proteins (PVX_112670 and PVX_110810A). In the Brazilian cohort, all IgM + RBP2b IgG performed (slightly) better than RBP2b IgG alone, with the exception of PVX_110810A. The best combination was PVX_087885 IgM + RBP2b IgG (AUC 0.88) for Thailand and PVX_082735 IgM + RBP2b IgG (AUC 0.85) for Brazil.

## Discussion

Serological markers of recent exposure to *P. vivax* infections could play an important role in malaria elimination by delineating areas of ongoing transmission and identifying individuals with a high chance of carrying hypnozoites in their livers. In this study we aimed to improve upon an existing set of serological exposure markers by incorporating IgM, in addition to IgG, responses against these proteins. We demonstrate that two proteins in particular, PVX_087885 (RAMA) and PVX_082735 (TRAP), induce IgM responses reflective of recent exposure to *P. vivax* within the past 9 months in endemic regions of Thailand and Brazil. They have similar accuracy as compared to these same antigens using IgG (RAMA performs slightly better with IgG, TRAP performs slightly better with IgM). However, the accuracy of these classifications overall is poorer than for the top performing serological marker (RBP2b) when using IgG, and generally the IgM AUC values for each antigen were lower than the corresponding IgG AUC values (Table S2). The poorer performance of IgM responses against these proteins than IgG likely relates to the acquisition and maintenance of IgM antibody responses following *P. vivax* infections. IgM is expected to be short-lived following infection, and supporting this we find the classification accuracy does improve if we define recent exposure within a shorter time frame (i.e. 1-6 months rather than 9). Another contributing factor is likely the high background in the non-malaria exposed controls for IgM (Figure S1 and S2), compared to our previous results for IgG [3].

When we combined each of the IgM responses against the 18-20 *P. vivax* proteins tested with the IgG response against RBP2b we demonstrated a clear improvement in classification performance compared to the single-antigen IgM response alone. However, there was only a slight improvement compared to the single-antigen RBP2b IgG alone, signifying that incorporation of IgM responses into the classification algorithm is unlikely to result in substantial improvements in classification. A limitation of our research is that we did not exhaustively test all combinations of IgG and IgM against the down-selected panel of 18-20 *P. vivax* proteins. We also only measured IgM responses against 18-20 of the top *P. vivax* proteins as indicated by their classification performance when using IgG responses; an alternate approach would have been to measure IgM responses to the full panel of 60 proteins. However, we have already demonstrated that incorporating more than 5 IgG responses results in only marginal improvements in classification performance compared to RBP2b IgG alone, and thus this approach (of exhausting all options) is unlikely to yield better results.

Finally, we ultimately aim to develop a point-of-contact test to be used in the field. It would be a more complicated and costly test if both IgG and IgM responses were required to be measured. We will therefore not be pursuing IgM responses in our optimization of our novel panel of serological exposure markers, and will instead focus on other avenues for improved performance of signals we can obtain from the IgG responses.

## Data Availability

Data are available within the manuscript and supplementary files.

## Acknowledgments

We gratefully acknowledge the extensive field teams that contributed to sample collection and qPCR assays, including Wang Nguitragool, Andrea Kuehn, Yi Wan Quah, Piyarat Sripoorote, and Andrea Waltmann. We thank all the individuals that participated in each of the studies, and thank the Australian and Thai Red Cross for donation of plasma samples. We thank the Volunteer Blood Donor Registry at WEHI for donation of plasma samples, and Lina Laskos and Jenni Harris for their collection and advice. We thank Christopher King (Case Western Reserve University) for provision of the PNG control plasma pool. We thank Fumie Matsuura (CellFree Sciences) and Christele Huon (Institut Pasteur) for contributing to expression of proteins.

## Footnotes

(1) RJL, MW, TT and IM are inventors on patent PCT/US17/67926 on a system, method, apparatus and diagnostic test for *Plasmodium vivax*. No other authors declare a conflict of interest.

(3) Part of this work was presented by RJL at the 47^th^ Annual Scientific Meeting of the Australasian Society for Immunology, December 2018, Perth, Australia.

(4) Correspondence and requests for reprints should be addressed to Professor Ivo Mueller, The Walter and Eliza Hall Institute of Medical Research, 1G Royal Parade, Parkville 3052, Victoria, Australia, +61 3 9345 2936.

